# The Impact of COVID-19 on the Management of Heart Failure -A United Kingdom Patient Questionnaire Study

**DOI:** 10.1101/2020.10.03.20205328

**Authors:** R Sankaranarayanan, N Hartshorne-Evans, S Redmond-Lyon, J Wilson, H Essa, A Gray, L Clayton, C Barton, FZ Ahmed, C Cunnington, DK Satchithananda, C Murphy

**Author notes:** (Corresponding Author) Email –.

## Abstract

**Aim:** The coronavirus disease 2019 (COVID-19) pandemic has created significant challenges to healthcare globally, necessitating rapid restructuring of service provision. This questionnaire survey was conducted amongst adult heart failure (HF) patients in the United Kingdom (UK), to understand the impact of COVID-19 upon HF services.

**Methods and Results:** The survey was conducted by the Pumping Marvellous Foundation (PMF), a UK HF patient charity. “Survey Monkey” was used to disseminate the questionnaire in the PMF’s online patient group and in 10 UK hospitals (out-patient hospital and community HF clinics). 1050 responses were collected (693/1050-66% women); 55% (579/1050) were aged over 60 years. Anxiety level was significantly higher regarding COVID19 (mean 7±2.5 on anxiety scale of 0 to 10) compared to anxiety regarding HF (6.1±2.4; p<0.001). Anxiety was higher amongst patients aged ≤60 years about HF (6.3±2.2 versus 5.9±2.5 in those aged >60 years; p=0.005) and COVID-19 (7.3±2.3 versus 6.7±2.6 those aged >60 years; p<0.001). 65% respondents (686/1050) reported disruption to HF appointments (cancellation or postponement) during the lockdown period. 37% reported disruption to medication prescription services and 34% reported inability to access their HF teams promptly. 32% expressed reluctance to attend hospital (25% stated they would only attend hospital if there was no alternative and 7% stated that they would not attend hospital at all).

**Conclusions:** The COVID-19 pandemic has caused significant anxiety amongst HF patients regarding COVID-19 and HF. Cancellation or postponement of scheduled clinic appointments, investigations, procedures, prescription and monitoring services were implicated as sources of anxiety.

## Introduction

Coronavirus disease 2019 (COVID-19) is caused by severe acute respiratory syndrome coronavirus 2 (SARS-CoV-2) (1). COVID-19 evolved into an international public health crisis rapidly after being discovered in December 2019, and was declared a pandemic by the World Health Organisation on March 11, 2020 (2). In the United Kingdom (UK) the accelerating pace of the COVID-19 pandemic resulted in the British government issuing a public lockdown from the 23^rd^ of March (3) and a rapid shift in the focus of care in the National Health Service (NHS) ensued. Lockdown measures were first eased in England from May 13, 2020 (4), whereas other devolved UK nations eased lockdown measures more slowly (for example Scotland from May 28^th^ onwards).

The COVID-19 pandemic has created an unprecedented challenge to healthcare resources throughout the world. Hospitals rapidly restructured services in order to enable them to cope with the predicted high numbers of COVID-19 patients. This led to the cancellation or postponement of elective appointments, and both inpatient as well as outpatient services were restricted to prioritise urgent cases. In particular, elective face to face appointments were reduced significantly and the threshold for admitting patients increased (5) in order to minimise the risk of exposure to COVID-19 to at-risk groups such as those with underlying cardiovascular conditions (6). In addition to this, patients have also been reluctant to attend hospital due to self-isolation, fear of contracting infection and also perhaps a mis-interpretation of government guidance that healthcare services are not open for non-COVID-19 healthcare including emergencies (7). This trend has been reflected in the significantly reduced hospitalisation rates for several acute cardiovascular conditions (8) such as acute coronary syndrome (9-11) as well as acute HF during the ongoing pandemic in the UK (12, 13) but also in other parts of the world (14, 15), with these results suggesting a 50% reduction in acute HF hospitalisations during the first wave of the pandemic. HF patients also received seemingly conflicting advice, with initial communication from Public Health England (PHE) advising that they were amongst those at increased risk of severe illness from COVID-19 (16), however subsequent communication excluded HF patients from the list of patients with chronic conditions classed as “clinically extremely vulnerable” (17).

The primary aim of this HF patient study was to ascertain the impact of the COVID-19 lockdown upon adult HF patient care. The pandemic response has also prompted a rapid evolution of service delivery such as the use of virtual clinics (using telephone/video technology), remote monitoring and ambulatory care. The Heart Failure Society of America and the European Society of Cardiology both recommend the use of tele-health strategies such as virtual clinics (remote consultation using telephone or video technology) and remote monitoring of patient health parameters (weight, pulse, blood pressure) as alternatives to minimise the need for in-hospital visits (18, 19) and recent evidence during the pandemic has also suggested that telemedicine can have beneficial outcomes in HF (5, 20). However, it is currently unclear whether the full spectrum of service reconfiguration is acceptable as well as accessible to patients and there is currently a lack of feedback regarding patients’ opinions about suitability of various tele-health strategies. We therefore incorporated patient feedback regarding service innovations into our survey and also sought to gauge HF patients’ preferences regarding the newer models of care that they would like to see incorporated into or continued in their local HF services.

## Methods

The survey was conducted by the Pumping Marvellous Foundation (PMF) which is the UK’s largest patient-led heart failure charity. PMF is funded via donations and fundraising by individuals, with support from the NHS and charitable organisations together with corporate sponsorship. A HF Patient Survey Working Group was constituted in May 2020, consisting of 3 HF patients, 6 HF cardiologists, 1 HF advanced nurse practitioner and 1 HF nurse consultant. The HF patient survey working group devised the survey questionnaire (supplementary material).

The anonymous questionnaire survey was disseminated from June 15^th^ 2020 to Aug 10, 2020 to approximately 7000 adult HF patients. One mode of dissemination was through the online survey tool Survey Monkey, on the Pumping Marvellous Closed Online Patient Group. The other was by handing out 4000 cards (containing Quick Response (QR) codes with links to the survey) to HF patients who attended out-patient clinics or community heart failure clinics affiliated to 10 participating hospitals in the UK.

HF patients were asked to rate their anxiety about HF and COVID19 on a scale of 0 (no anxiety) to 10 (highest anxiety level). The survey also consisted of questions regarding the impact of COVID-19 lockdown upon out-patient HF clinic appointments (hospital as well as community HF services), cardiac investigations, procedures and access to medications as well as HF counselling. Opinions were also sought regarding preference for service changes enforced by the pandemic. Statistical comparisons were performed based on gender and age groups (patients aged up to 60 years versus those aged more than 60 years).

### Statistical Analysis

For the descriptive statistics of our patient population, we represent continuous variables as means with standard deviations (mean ± SD) when normally distributed and compared using a Student’s T test. For non-normal data, the parameters are described using medians with interquartile ranges and compared using Mann –Whitney test. Categorical data are expressed as percentages and compared using the chi-square test.

## Results

A total of 1050 adult heart failure patients and carers were surveyed from England (80%, 840) Scotland (10%, 100), Wales (8%, 87) and Northern Ireland (2%, 23). The majority of survey respondents (696 out of 1050 -66%) were female and 55% (579/1050) were aged more than 60 years. Comprehensive patient demographics are detailed in Table 1.

**Table 1.** Demographics of HF Questionnaire Survey Respondents

| **Characteristic** | **n =1050** | **Percent (%)** |
| --- | --- | --- |
| **Gender** |  |  |
| Male | 357 | 34.4% |
| Female | 693 | 66.6% |
| Prefer not to say | 5 | 0.4% |
| **Age** |  |  |
| 18-30 | 8 | 1% |
| 31-40 | 38 | 3% |
| 41-50 | 132 | 12% |
| 51-60 | 293 | 28% |
| 61-70 | 309 | 29% |
| 71-80 | 215 | 21% |
| >80 | 55 | 5% |
| **Identification** |  |  |
| Patient living with heart failure | 950 | 87% |
| Carer of patient | 100 | 9% |
| **Location** |  |  |
| England | 840 | 80% |
| Scotland | 100 | 10% |
| Wales | 87 | 8% |
| Northern Ireland | 23 | 2% |

The survey respondents reported a significantly higher degree of anxiety regarding COVID19 in comparison to anxiety regarding HF itself (mean 7 ±2.5 on the anxiety scale versus 6.1 ± 2.4; p<0.001). This trend was similar amongst patient sub-groups based on gender (males HF anxiety score 6.0 ± 2.2 vs. COVID anxiety score 7 ±2.5; p<0.001 and females HF anxiety score 6.1 ± 2.4 vs. COVID anxiety score 7 ± 2.4; p<0.001) as well as age groups i.e., patients aged up to 60 years versus those aged more than 60 years.

65% (686/1050) of all respondents reported that their heart failure appointments were negatively impacted during the lockdown period of the COVID19 pandemic with 43% cancellation of scheduled hospital clinic appointments (321/743 appointments) and 22% cancellation of community HF clinic appointments (114/523) (Figure 1). Cardiac investigations (echocardiogram, holter monitoring, cardiac magnetic resonance imaging, cardiac computed tomography scans) or procedures (angioplasty, pacemaker, cardiac bypass operation and radiofrequency ablation) were also affected with 37% (362 out of 994) appointments cancelledand46% (460/994) postponed (Figure 2). As shown in figure 2, the most commonly cancelled investigation was cardiac MRI (112 reported cancellations) and the most commonly cancelled procedure was pacemaker implantation (38 cancellations). The most commonly postponed investigation was the holter monitor test (101), and procedure was pacemaker implantation (60 postponements). The most frequently performed investigation or procedure during this period was echocardiography (117). 29% (305/1050) of respondents reported that their appointments for HF counselling were cancelled.

**Figure 1.**
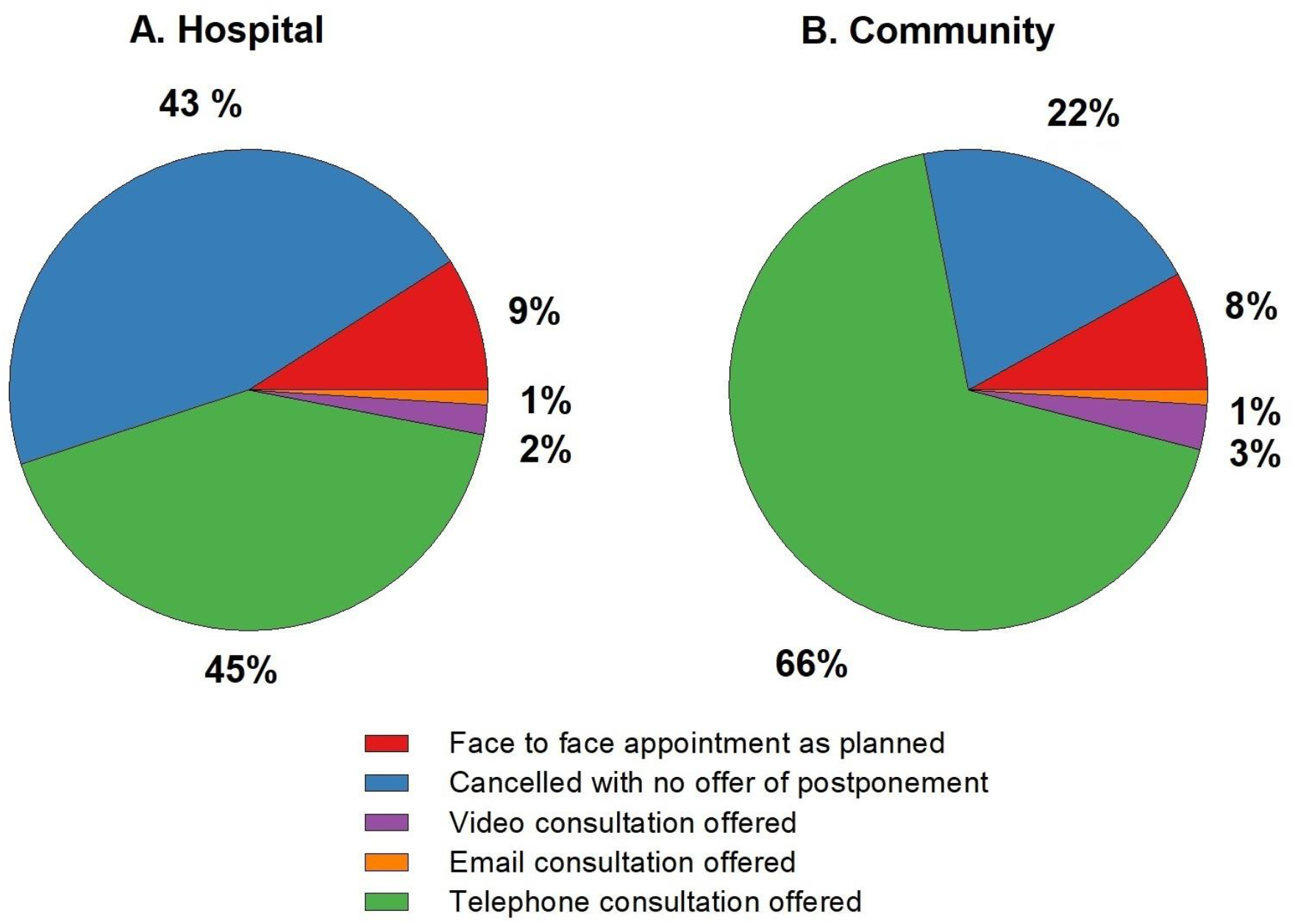
Outcomes of scheduled heart failure appointments during the lockdown period.

**Figure 2.**
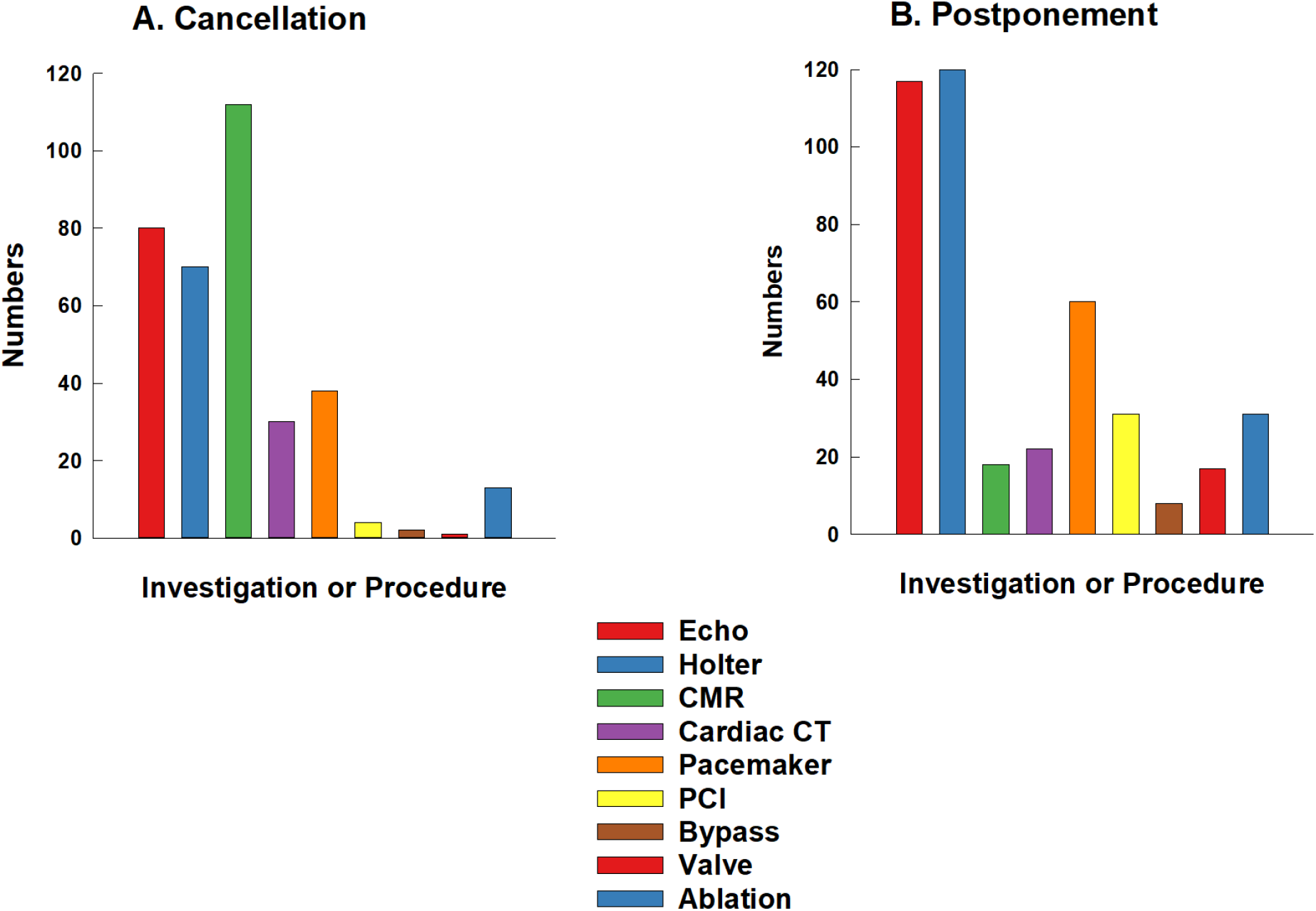
Number of investigations or procedures adversely affected.

A. Cancellation B. Postponement

37% (389/1050) of HF patients reported that their medication prescription was adversely affected by the pandemic and 34% (357/1050) reported that they could not access their HF services/teams promptly. The majority of patients (68%) reported a willingness to attend hospital appointments if required (18% responded that they would have no concerns attending hospital and 50% stated that they would attend hospital if there were appropriate measures to ensure patient safety, such as social distancing, availability of masks, hand-sanitisation facilities). However, nearly a third of HF patients surveyed (32%) reported a reluctance to attend hospital (25% stated that they would only attend hospital if there was no alternative and 7% stated that they would not attend hospital at all).

Respondents were also asked to choose service changes or new models of care that they would prefer to be continued or to be introduced in their local HF services (Figure 4). An overwhelming majority (71%) chose the “One stop diagnostic HF clinics”, consisting of a single visit incorporating HF consultant review and echocardiography. With regards to alternatives to physical attendance at HF clinics, telephone consultations were more popular (52% preferred this option) in comparison to video consultation (preferred by 34%). 36% of HF patients stated that they would prefer home intravenous diuretic therapy as an alternative to hospital admission and 29% preferred ambulatory or outpatient intravenous treatment. 51% of patients preferred to have their blood pressure, pulse and weight monitored at home (“home-monitoring”).

**Figure 3.**
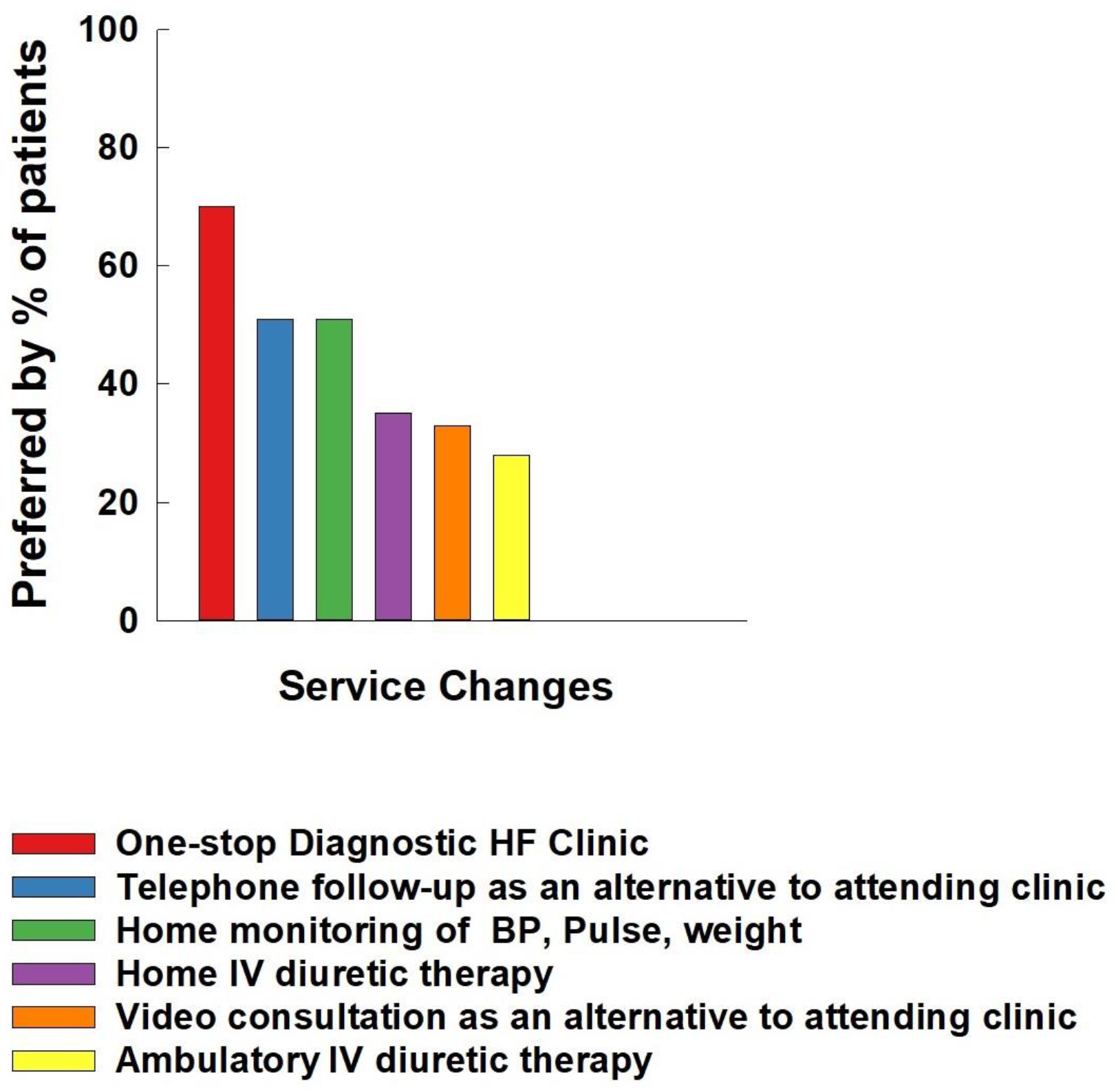
Services chosen by heart failure patients as preferred to be continued or introduced.

**Figure 4.**
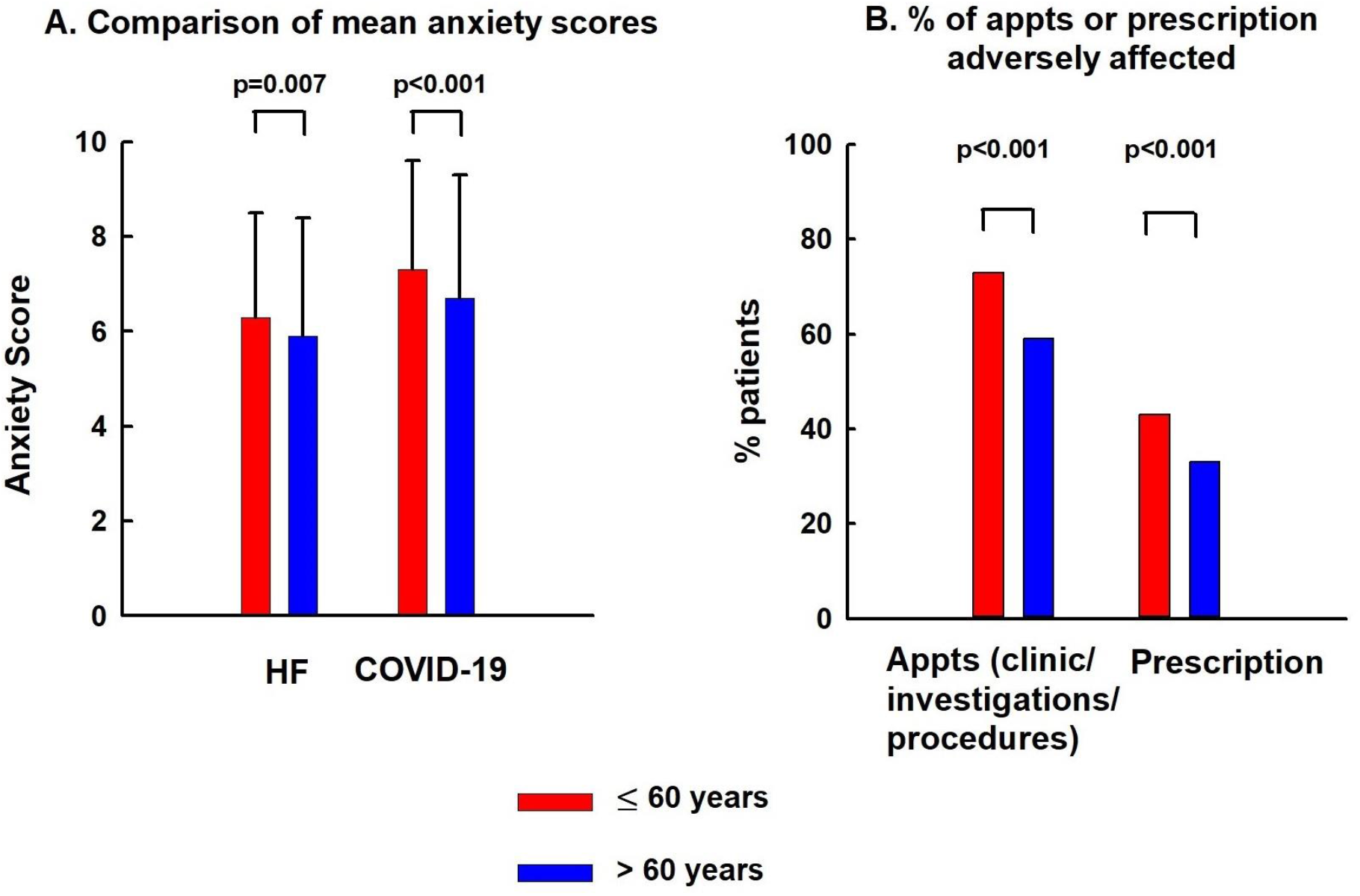
Age group comparisons (HF patients aged ≤60 years old versus those aged >60 years old)

Survey responses were further analysed based on age groups (age ≤60 years versus > 60 years) and this is illustrated in detail in Table 2. There was a greater degree of anxiety amongst younger patients (aged ≤60 years) versus older patients (aged>60 years) both for HF (6.3 ± 2.2 vs. 5.9 ± 2.5; p=0.005) as well as for COVID-19 (7.3 ± 2.3 vs. 6. 7± 2.6; p=0.0003). Whilst HF patients aged ≤60 years had a higher number of appointments scheduled (2.5 ± 1.6) compared to older patients (1.8 ± 1.2; p<0.001), younger patients also reported a higher proportion of disruption (cancellations or postponement) to clinic appointments as well as appointments for investigations/procedures in comparison to older patients (73% of patients in the group ≤60 years vs. 59% in the group aged >60 years; p<0.001). It is therefore possible that the higher degree of anxiety was influenced by the fact that patients aged≤ 60 years experienced a significantly higher proportion of appointment cancellations (hospital and community) as well as disruptions to medication prescription or monitoring. Older patients (age>60 years) were extremely wary of attending hospital with 34% opting to only attend hospital if there was no alternative or to not attend hospital at all, in comparison to 26% of patients aged ≤60 years who expressed this view (p=0.004).

**Table 2:** Survey analysis based on age groups (up to 60 years old versus over 60 years old)

|  | **≤60 years**<br>n=471/1050 | **>60 years**<br>n=579/1050 |
| --- | --- | --- |
| **Anxiety about HF**(Mean ± SD)<br>(0=no anxiety and 10=highest anxiety level) | 6.3 ± 2.2 | 5.9 ± 2.5 (p=0.005) |
| **Anxiety about COVID**(Mean ± SD)<br>(0=no anxiety and 10=highest anxiety level) | 7.3 ± 2.3<br>HF vs COVID<br>(p<0.001) | 6.7 ± 2.6 (p=0.0003)<br>HF vs COVID<br>(p<0.001) |
| **Number of appointments scheduled**<br>(Mean ± SD) | 2.5 ± 1.6<br>(1179<br>appointments) | 1.8 ± 1.2<br>(1081 appointments)<br>p<0.001 |
| **% of patients with appointments,**<br>**investigations or procedures adversely**<br>**affected** | 73% of patients<br>(344/471) | 59% of patients<br>(342/579); p<0.001 |
| Face-to face hospital appointments cancelled<br>(% of total appointments) | 48% (190/393) | 37% (131/350); p =0.003 |
| Community appointments cancelled<br>(% of total appointments) | 28% (64/228) | 17% (50/295); p=0.002 |
| Investigation /procedure cancelled<br>(% of total appointments) | 40% (221/558) | 32% (141 /436); p=0.04 |
| Investigation/procedure postponed<br>(% of total appointments) | 51% (279/558) | 41% (181/436); p=0.008 |
| **Prescription or monitoring affected**<br>(% of patients) | 43% (204/471) | 33% (189/579) ;p<0.001 |
| **“Don’t feel can access services”**<br>(% of patients) | 33% (155/471) | 34% (199/579); p =NS |
| **Patients willing to attend hospital if required**<br>(% of patients) | 73% (345/471) | 66% (381/579) ;p=0.01 |
| “I would have no concerns about attending the<br>hospital | 17% (79/471) | 19% (108/571) |
| “I would attend the hospital if there were<br>appropriate measures in place to ensure my<br>safety” | 56% (266/471) | 47% (273/579) |
| **Patients reluctant to attend hospital** | 26% (126/471) | 34% (198/579); p=0.004 |
| “Would attend hospital only if no alternative” | 23% (110/471) | 26% (150/579) |
| “Would never attend hospital” | 3% (16/471) | 8% (48/579) |
| **Preference for new service models** |  |  |
| Telephone appointment | 56% (264/471) | 49% (282/579) ;p=0.02 |
| Video consultation | 44% (206/471) | 26% (149/579); p<0.001 |
| Ambulatory IV diuretic | 33% (156/471) | 25% (147/579);p=0.005 |
| Home IV diuretic | 40% (187/471) | 33% (191/579);p=0.02 |
| Home monitoring | 56% (265/471) | 47% (270/579);p=0.003 |
| 1 stop HF diagnostic clinic | 75% (354/471) | 67% (387/579); p<0.001 |
| Patients not under care of hospital HF team | 10% | 25% |
| Patients not under care of community HF team | 29% | 34% |

There was also a significant difference in terms of preference for newer models of HF services between the two age groups compared, with these service changes seemingly less popular amongst older HF patients (aged >60 years).

## Discussion

Our questionnaire-based adult HF patient survey is the first reported study which describes the impact of the COVID-19 pandemic lockdown period upon HF services from a patient perspective in all four devolved nations in the UK. Our results show high anxiety levels particularly regarding COVID-19 and especially amongst younger patients. It is likely that this was exacerbated by the cancellation or postponement of a large proportion of clinic appointments (hospital as well as community), elective investigations and procedures, with the majority of survey respondents (65%) reporting that their HF appointments were adversely affected through cancellation or postponement. Whilst reassuringly the majority of respondents surveyed reported that they felt able to access their HF services, it is of great concern that over a third of patients reported that they had difficulties accessing their local HF services or obtaining HF medications.

A number of studies have shown a significant reduction in admissions due to HF during the 1^st^ peak of COVID-19 (12, 14, 21). As demonstrated by our study, possible reasons for this include a reluctance or unwillingness to attend hospital (this was seen especially amongst the older patient group aged>60 years in our study), due to anxiety amongst HF patients about contracting COVID-19 in hospital as well as a perception that clinical services were only open to treat COVID-19 cases. This is reflected in our survey responses which showed higher anxiety levels regarding COVID-19 in comparison to anxiety about HF. High anxiety levels have been independently correlated with adverse HF outcomes in previous studies (22). Interestingly despite older people having a predilection for more severe COVID-19 illness, anxiety levels were greater amongst the younger group of patients (aged up to 60 years). This is contrary to the common perception that older patients are likely to experience more anxiety due to a combination of social isolation as well as fear of contracting severe COVID-19 illness. This paradoxical result may be in part due to the greater degree of disruption to clinic appointments and monitoring, as well as interruption to HF medication prescriptions amongst younger patients. In addition, it is also possible that younger patients feared hospitalisation or dying more than older people and were perturbed about the possible impact of COVID-19 upon their family or dependents such as school age children. Other possible reasons for greater apprehension amongst younger HF patients could have been related to specific financial concerns, fear of being made unemployed or feeling pressurised to work in environments where physical distancing was difficult to achieve.

Patients with a new diagnosis of HF typically undergo an armamentarium of diagnostic investigations, followed by appropriate guideline directed management through timely follow-up. Cancellation or postponement of these appointments can lead to HF decompensation, worsening of quality of life and also increase patients’ anxiety levels regarding accessibility of HF services, risk of hospitalisation or dying prematurely thereby triggering a vicious cycle. Delays in optimisation of prognostic HF medications, investigations and procedural treatments, are likely to lead to worse HF outcomes such as acute decompensation, worsening of quality of life and increased mortality.

Significantly, around one third of HF patients in our survey expressed a reluctance to attend hospital (25% stated that they would only attend hospital if there was no alternative and 7% stated that they would not attend hospital at all). The prevalence of this viewpoint was significantly higher amongst the older patient group (aged >60 years). Mixed messages regarding isolation or shielding may have caused confusion, leading to indecision amongst HF patients about accessing healthcare services. This may have contributed to the significantly lower hospital admission rates for HF during the pandemic (12, 13), but also to the worse outcomes in the form of higher mortality (13). It is crucial therefore that healthcare services reach out to all patients, with reassurances that adequate precautions are being taken in healthcare settings (such as physical distancing, use of hand-washing/sanitising facilities and personal protective equipment), in order to minimise the risk of transmission of coronavirus to patients in healthcare settings.

The response to the COVID-19 pandemic triggered a rapid restructuring of healthcare services with an increased use of and reliance on tele-cardiology services. The data available on the tele-management of cardiology patients is conflicting. Whilst some trials have shown beneficial end points (23, 24), others have suggested a lack of improvement in outcomes (25, 26). Our survey results offer the first published report of adult HF patients’ opinions regarding the acceptance and preference for ongoing continuation of these newer service models. Overall, the ‘one-stop diagnostic HF clinic’ was the most popular service preference with over 70% of respondents opting for it to be continued. This concept minimises the need for repeat HF clinic attendances by incorporating echocardiography and HF specialist review during the same consultation, thereby improving patient convenience and possibly reducing risk of exposure to COVID by minimising hospital/healthcare facility attendance. This is also the objective of other services such home monitoring (blood pressure, pulse, weight), home treatment or ambulatory treatment with intravenous diuretics and virtual clinics (using telephone or video technology). However, these newer service models appeared to be less popular with patients, particularly with older patients. This is a pivotal finding of the study, with possible far-reaching consequences because HF patients are frequently elderly and have a significant number of existing co-morbidities (27).Virtual or remote consultations can be challenging for health care providers to deliver due to the inability to conduct a physical examination, communication barriers with patients who have hearing impairment, reduced opportunity for consultation input from carers and language barriers. Potential solutions include the use of tele-health home monitoring of parameters such as pulse, blood pressure, heart rate and weight, educating patients to self-examine for the presence/absence of fluid overload and use of multi-participant teleconference technology or loudspeaker function at the patient facing side of the consultation to help include carers and interpreters in the consultation process. It is therefore crucial that HF services engage with their patients to co-design and evaluate novel services prior to widespread implementation. This should include a comprehensive understanding of: patient access barriers; acceptability of novel services; patient preferences; and local geographic and socio-economic challenges that may exist, so that patient care can be delivered with parity.

To date, this is the 1^st^ reported HF patient survey that explores the direct impact of COVID-19 upon adult HF patient care, from a patient perspective. This study has several strengths including but not limited to a large sample size distributed across 10 hospitals in the UK with inclusion of a wide range of ages from all four nations in the UK. The study also benefits from having been undertaken during the final phases of the UK lockdown. This allowed us to capture a more complete picture of any disruptions that HF patients may have experienced during this time period. Importantly, the results should inform adult HF service transformation and resilience strategies in the event of a second pandemic wave, including the adoption of an approach that maintains elective services for people whose underlying health condition places them at high risk of hospitalisation or premature death. Such an approach has the potential to minimise the anxiety and disruption to care, experienced by HF patients.

## Limitations

General limitations exist when using a survey questionnaire study model. Such limitations are applicable to this study including the potential for responder bias and a response rate of approximately 15%. It is possible that non-responders experienced no disruption to their appointments during the pandemic or conversely, that they were either unable to access the technology to complete the survey or were not aware of the survey due to lack of contact with their HF team.

## Conclusion

The COVID-19 pandemic represents a unique inflection point in healthcare provision and required a rapid response from healthcare systems in order to cope with the burgeoning demand created by a new multisystem condition. We are already experiencing further waves of this pandemic across several geographical areas. Our survey results highlight that patients with chronic health conditions such as heart failure need to feel reassured that health care services can respond in a timely and robust manner when treating them, whilst also being able to cope with the burden created by COVID-19.It is likely that the resulting healthcare disruptions for non-COVID conditions in multiple countries, led to the public concluding that healthcare facilities were “COVID-19 Hospitals” and therefore less able to look after them. This, in combination with a real-world reduction in services offered, possibly resulted in patients feeling that they could not rely on the services they had previously relied upon and had confidence in. It is important that healthcare authorities regain the trust of patients with chronic conditions such as HF, through firm reassurance that healthcare services are ready and capable of coping with the extra demand created by COVID-19. However, this requires to be balanced with competing demands on other services. The results of our nationwide HF patient survey should influence the future direction of adult HF care in the event of future waves of COVID-19 and help optimise service delivery under these exceptional circumstances, in order to minimise disruption to patient care.

## Ethics

Our study complies with the Declaration of Helsinki, the research protocol complied with the institutional ethics and written patient consent was waived due to complete anonymization of data at all stages of the study from the time of responses to the questionnaire survey by patients as well as analysis.

## Data Availability

Raw data were generated at the Pumping Marvellous Foundation by analysis of data from https://www.surveymonkey.co.uk/r/K2DSBPZ. Derived data supporting the findings of this study are available from the corresponding author [RS] on request

## Acknowledgements

nil

## Funding

no external sources of funding were used

## Conflicts of Interest

none declared

## HEART FAILURE PATIENT SURVEY REGARDING THE IMPACT OF COVID-19 UPON HEART FAILURE PATIENT CARE

Q.1 Are you *(select one option)*
  [1] Male
  [2] Female
  [3] Prefer not to say
  GO TO Q.2
Q.2 How old are you? *(select one option)*
  [1] 18-30
  [2] 31-40
  [3] 41-50
  [4] 51-60
  [5] 61-70
  [6] 71-80
  [7] >80
  GO TO Q.3
Q.3 Where do you live? *(select one option)*
  [1] England
  [2] Scotland
  [3] Wales
  [4] Northern Ireland
  GO TO Q.4
Q.4 Anxiety - On a scale of 0 (no anxiety) to 10 (high anxiety level), how do you rate your anxiety level about
  [a] Heart Failure 0 1 2 3 4 5 6 7 8 9 10
  [b] COVID-19 0 1 2 3 4 5 6 7 8 9 10
  GO TO Q.5
Q.5 Specifically related to the **managment of your heart failure**,how many appointments were scheduled in the last 12 weeks (since the outbreak of the COVID-19 pandemic)? 0 1 2 3 4 5 6 7 8 9 10 Please select which of the following appoinments/procedures, if any, were scheduled in the last 12 weeks (since the outbreak of the COVID-19 pandemic) *Tick all that apply*
  [1] Scheduled hospital appointment with your heart failure team
  [2] Scheduled community appointment with your heart failure team (e.g heart failure nurse to visit you at home, at community based clinic)
  [3] Heart failure counselling
  [4] Cardiac investigations (e.g echocardiogram, heart monitor)
  [5] Cardiac procedures/operations
  [6] None of the above
  IF CODE 1,2,3,4,5 AT Q.5 GO TO Q6. IF CODE 6 AT Q.5 GO TO Q8
Q.6 Have any of your scheduled **heart failure appointment/s, investigations/operations** been affected by the ongoing COVID-19 pandemic? YES GO TO Q7 NO GO TO Q8
Q.7 What impact has the ongoing COVID-19 pandemic had on your heart failure team appointment/s, cardiac investigations/procedures /operations?
  **A.CANCELLATION**
    A.1 **Hospital heart failure team appointments was cancelled (if applicable)** Alternative to hospital appointment was offered *select one option only:*
      1 Email
      2 Telephone consultation
      3 Video consultation
    A.2 **Community heart failure team appointments was cancelled (if applicable)** Alternative to community appointment was offered *select one option only:*
      1 Email
      2 Telephone consultation
      3 Video consultation
    A.3 **Heart failure counselling was cancelled (if applicable)** Alternative to Heart failure counselling was offered *select one option only*
      1 E-mail
      2 Telephone consultation
      3 Video consultation
    A.4 **Scheduled Cardiac Investigations Cancelled (if applicable)** *select all that apply*
      1 Echocardiogram cancelled
      2 Heart Monitor Test cancelled
      3 Cardiac MRI cancelled
      4 Cardiac CT cancelled
    A.5 **Cardiac procedure/operation Cancelled (if applicable)** *select all that apply*
      1 Pacemaker cancelled
      2 Angioplasty cancelled
      3 Heart Bypass cancelled
      4 Valve Procedure cancelled
      5 Ablation cancelled
  **B.POSTPONEMENT**
    B.1 **Hospital heart failure team appointments was postponed (if applicable)** Alternative to hospital appointment was offered *select one option only*
      1 Email
      2 Telephone consultation
      3 Video consultation
    A.2 **Community heart failure team appointments was postponed (if applicable)** Alternative to community appointment was offered *select one option only:*
      1 Email
      2 Telephone consultation
      3 Video consultation
    A.3 **Heart failure counselling was postponed (if applicable)** Alternative to Heart failure counselling was offered *select one option only*
      1 E-mail
      2 Telephone consultation
      3 Video consultation
    A.4 **Scheduled Cardiac Investigations Postponed (if applicable)** *select all that apply*
      1 Echocardiogram postponed
      2 Heart Monitor Test postponed
      3 Cardiac MRI postponed
      4 Cardiac CT postponed
    A.5 **Cardiac procedure/operation Postponed (if applicable)** *select all that apply*
      1. Pacemaker postponed
      2. Angioplasty postponed
      3. Heart Bypass postponed
      4. Valve Procedure postponed
      5. Ablation postponed
      GO TO Q8
    Q.8 Has the **prescription /monitoring** of your heart failure **medication** been affected by the ongoing COVID-19 pandemic? YES *(select all that apply)*
      [1] Medication changes postponed
      [2] Intravenous therapy postponed
      [3] Difficulty obtaining repeat prescriptions
      [4] Difficulty having blood tests checked
      GO TO Q9 NO GO TO Q.9
    Q.9 Do you feel you can access your heart failure services promptly if your symptoms worsen? YES
      - Are they responsive? *(select one option)*
        [1] Yes
        [2] No GO TO Q10
        [3] NO
        GO TO Q10
    Q.10 If you were offered a hospital outpatient consultation during the COVID-19 pandemic, how willing would you be to attend the hospital ? *select one option*
      [1] I would have no concerns about attending the hospital
      [2] would attend the hospital if there were appropriate measures in place to ensure my safety (i.e. appropriate social distancing, availability of masks/hand washing facilities etc)
      [3] I would attend the hospital only if there was no alternative
      [4] I would not attend the hospital under any circumstances GO TO Q11
    Q.11 Which of these service changes or new models of care would you like to see established/continue in your area? *select all that apply*
      [1] Telephone follow-up (as an alternative to attending an outpatient clinic)
      [2] Video consultation (as an alternative to attending an outpatient clinic)
      [3] Ambulatory intravenous diuretic therapy (injections to remove excess fluid), i.e. daily visit to a hospital outpatient unit (as an alternative to hospital admission for worsening heart failure)
      [4] Home intravenous diuretic therapy (injections to remove excess fluid), i.e. daily visit from a nurse to give intravenous therapy at home (as an alternative to hospital admission for worsening heart failure)
      [5] Home monitoring of blood pressure, pulse, weight
      [6] One stop clinics - i.e. single visit consisting of echocardiogram, consultant and heart failure nurse specialist review

THANK YOU!

## Notes

### Competing Interest Statement

The authors have declared no competing interest.

### Funding Statement

No external funding was received

### Author Declarations

All necessary ethical guidelines have been followed. Institutional ethics approval obtained from the advisory board of the Patient-Led Charity Pumping Marvellous Foundation and the audit and research department at Liverpool University Hospitals NHS Foundation Trust

